# Health Care Student Orientations to Giving and Using Feedback in Workplace Learning: The Dual-role Feedback Orientation Scale

**DOI:** 10.1101/2025.11.12.25339987

**Authors:** Renske A.M. de Kleijn, Claudia J.M. Tielemans

## Abstract

Health professions education should prepare students to engage in feedback dialogues at the workplace. Studies often focus on students either giving or using feedback, and an instrument to address both dialogical roles is not yet available. Therefore, we adapted and extended the Feedback Orientation Scale into the Dual-role Feedback Orientation Scale (DrFOS). The DrFOS comprised 30 items, with separate User and Giver scales for Utility, Accountability, and Self-efficacy. The questionnaire was completed by 537 undergraduate 4-5^th^ year medical and nursing students. Exploratory factor analyses showed that the User and Giver subscales could be meaningfully and reliably discerned. Students reported remarkably high User Utility scores, thus believe that feedback is indispensable to learn. Their Self-Efficacy as Givers was relatively low. Additional cluster analysis indicated that students had either high or low overall dual-role feedback orientations, or seemed to value using feedback but did not feel very accountable or competent either as giver or as user. The findings show that the DrFOS is worthwhile using in future research in health professions education. These studies could adopt longitudinal or intervention designs and explore the relations of Dual-role Feedback Orientation to other relevant outcome measures.

## Introduction

It is uncontested that feedback is an essential element of health professions education (Bing-You et al., 2017; Ramani et al., 2019a; Tripodi et al., 2021; Van Der Leeuw & Slootweg 2013), especially when learning in the clinical workplace is a substantial part of undergraduate education. In the past decade more and more attention has been given to a proactive role of students in feedback processes (e.g., Molloy et al., 2020; Noble et al., 2020). This proactive role can include: students actively processing and using feedback they receive (e.g., Pelgrim et al., 2013; Van Der Leeuw & Slootweg, 2013), students actively asking for, or seeking feedback (e.g., Crommelinck & Anseel 2013; De Kleijn, 2023; Ramani et al., 2019b) Tripodi et al., 2021), and students not only being feedback receivers, but also feedback givers for their peers, teachers and/or supervisors (e.g., Fluit et al., 2013; Olvet et al., 2021). (Team)work in an authentic and complex clinical workplace requires flexible, bi-directional, *dialogical* communication. Such dialogues require two skillsets that together encompass all elements of the proactive role: (a) using feedback, which includes proactively seeking, understanding, and acting on feedback from a broad range of perspectives, and (b) giving feedback, proactively, to peers and superiors. We argue that healthcare education has the task to prepare students for both the role of feedback user, *and* that of feedback giver.

Only few empirical studies address healthcare students as prospective feedback givers to other health professionals. Ramani et al. (2019a), in their 12 tips for a feedback culture, do address the importance of health professionals’ roles of feedback provider and feedback recipient. In line with that, Tielemans et al. (2023a), presented their Westerveld feedback framework with seven criteria for interprofessional feedback dialogues, meaning that both the roles of feedback givers and users are described in light of these criteria. Note that they deliberately do not refer to feedback receivers or recipients, but feedback users. In their follow-up study they found that students struggled more with starting (interprofessional) feedback dialogues as a feedback giver, than with receiving or asking for feedback (Tielemans et al., 2023b). Building on the work of Ramani and Tielemans, research would need to explore to what extent attitudes towards giving and using feedback are related, naturally develop over time, and how they are affected by targeted interventions. But for future empirical studies to address student attitudes towards both using, and giving feedback at the workplace, it is essential that there are instruments available to do so. To fit this purpose, the present study aimed to adapt and expand an existing and validated feedback questionnaire: the Feedback Orientation Scale (FOS; Linderbaum & Levy, 2010).

Feedback orientation is defined as “an individual’s overall receptivity to feedback” (London & Smither, 2002, p81). In the context of performance and talent management, feedback orientation has been found to be associated with individual differences as well as organizational criteria like task performance and feedback seeking (Patel et al., 2019). In higher education, feedback orientations are found to be associated with feedback use (Winstone et al., 2021), goal-orientations (Winstone et al., 2021), self-assessment (Yan et al., 2020). Specifically, in health professions education, studies have found positive relations for students’ feedback orientations with performance measures (Chen et al., 2022; Rasheed et al., 2015), and with how students experience their supervisor feedback (Nolan & Loubier, 2018); Mills et al. (2023), found no differences between feedback orientations of medical students and internal medicine residents, and Tornwall and Ikonen (2024), showed that feedback orientations can be affected by educational interventions. They found that after a 7-week course, nursing students had increased feedback orientations (i.e. reported to be more receptive to feedback). In sum, the concept of feedback orientation has a strong conceptual foundation, is found to be related to relevant other concepts, and is applicable to settings of workplace learning, like in health professions education. However, given the current definition, it focuses only on the receiver, or user, role of feedback. We propose to extend it to also include an individual’s overall orientation towards giving feedback.

Feedback orientation is often measured with the Feedback Orientation Scale (FOS; Linderbaum & Levy, 2010). The FOS contains 20 items divided in 4 subscales: Utility, the belief that using feedback is instrumental in achieving goals /obtaining desired outcomes; Accountability, a sense of obligation to act on feedback; Self-efficacy, confidence in dealing with receiving feedback; Social awareness, the tendency to use feedback as to be aware of others’ views of oneself. With respect to Social awareness: low initial response rates required us to shorten the data collection instrument used in this study. As several educational authors have argued that social awareness is relatively less interesting in educational contexts (Kasch et al., 2022; Winstone et al., 2021), we did not include it in this study. We do use the other scales and extend them to a context of both receiving and giving feedback. We name the new instrument the Dual-role Feedback Orientation Scale (DrFOS).

In sum, given the need to study health care students’ and professionals’ perspectives on feedback user *and* giver skills, our research question was: to what extent can the DrFOS meaningfully measure and discern giver and user feedback orientations in clinical HPE?

## Method

### Participants

The participants were 5^th^ year medical and 4^th^ year nursing students at a medical and nursing school in the Netherlands. Every six weeks, a cohort of approximately 100 students, 30 5^th^ year undergraduate medical and 70 4^th^ year undergraduate nursing students, enroll in an obligatory two-day course on interprofessional feedback. For this course they return to the classroom, but all participants are in the workplace-learning phase of training and have been working in healthcare teams for at least a year. During this classroom-based course all students were in their workplace-based learning phase in their program. For a more detailed description of the course see Tielemans et al. (2023b).

### Procedure

In all cohorts from January 2022 to September 2023, at the start of the course, all students were invited to voluntarily complete an online questionnaire as part of their preparation for the first course day. Participation was voluntary and informed consent was gained before each questionnaire. In total 1,159 were invited to participate, and 611 students filled out the questionnaire. 74 students indicated that their data could not be used for research purposes which led to a final sample of 537 students and an estimated response rate of 46%. Ethical approval of this study was provided by the Dutch Association for Medical Education (NVMO), ERB file number: 2022.1.6.

### Instruments

In the Dual-role Feedback Orientation Scale, we include User Feedback Orientation and Giver Feedback Orientation. To measure User Feedback Orientation (UFO), we used the Utility, Accountability, and Self-efficacy items of the original Feedback Orientation Scale (Linderbaum & Levy, 2010). Small adaptations were done to fit the situation of teamwork in the clinical workplace; “at work” and “in a company” were replaced by “in health care practice” (item 1, 2, and 4). For two items “supervisor” was replaced with “team members” (item 4 and 9) and in three items we explicitly added the word “received” to feedback, so that the contrast with feedback given would be clearer (item 11, 12, and 13). Lastly, to better fit the context of students as opposed to graduated professionals, for item 12 we changed the word “others” to “peers”.

To measure Giver Feedback Orientation (GFO), the fifteen UFO items were mirrored to address the same topic from the perspective of a feedback giver (see online supplement). First, the initial English items for GFO were formulated in a group meeting with a communication teacher with a PhD in medical education and a full professor in medical education. Second, after this meeting all four team members individually finetuned the items in a shared document. Third, these items were discussed in a PhD meeting with twelve PhD students in (bio)medical education. Both authors decided on the final items, with the aim to stay as closely as possible to the formulation of the User items. For instance, the first item “Feedback contributes to my success in health care practice”, was mirrored to “Me giving feedback to team members contributes to their success in health care practice”. Fourth, two medical students filled out all thirty items while thinking aloud. This led to a few small changes in wording, but mainly resulted in not using the word “feedback user”, but “feedback receiver.” And even though that does not match the current ideas about proactivity in feedback^e.g.,13^, this did match better to the students’ ideas and jargon with respect to feedback. Fifth, after the items were finalized in English, CT translated them to Dutch. Using backward translation, a fellow PhD student translated the Dutch items back to English and based on that process, small changes were made to the wording of the items in Dutch.

### Analysis

First, we checked the data on missingness to see whether there were items that were left open substantially more often than others. Second, we explored to what extent user and giver feedback orientations could be meaningfully discerned. Therefore, as did Linderbaum and Levy (2010), several exploratory factor analyses (EFA) were run on the 30 questionnaire items. Following the recommendations of Costello and Osborne (2005), we used a maximum likelihood (ML) estimation and oblique rotation (Oblimin). We considered all factors with an eigenvalue of >1.00 and constrained the number of factors to six. Third, we conducted reliability analysis on the scales to see whether the scales would meet the criterium of Cronbach’s alpha being >.70. Fourth, to see to what extend the factors differed within students, scale means were computed and compared using repeated measures ANOVA, with Bonferoni posthoc tests. Lastly, to investigate the relations between the scales Pearson’s correlation coefficients were computed. We interpreted correlations of .10 to .30 as small, .30 to .50 as medium, .50 to .70 as large and >.70 as so high that the scales might not be measuring meaningfully different variables. Lastly, to explore patterns between individuals, we performed a two-step cluster analysis on the scale scores and inspected the cluster quality. In case of cluster quality being fair or good, the results are presented and interpreted.

## Results

### Missingness

Within the 537 completed questionnaires, missingness ranged from one to six missing values per item. This indicates that none of the items was omitted substantially more often than others. We interpret this to mean that none of the items were incomprehensible or not applicable to a lot of students.

### Exploratory factor analysis and Reliability analysis

Table 1 presents the means and standard deviations for all thirty items. An exploratory factor analysis including all factors with eigenvalues of >1.00 yielded seven factors. When constraining the factor solution to six factors, the factors clearly represented the intended subscales. Therefore, we decided to continue with the six-factor solution (see Table 1 for the factor loadings and item descriptives). The subscales User Utility, Giver Utility, User Self-efficacy, and Giver Self-efficacy are clearly represented by factor 3, 5, 1, and 2 respectively. For both Accountability scales, some cross loadings were found. More specifically for the User items, number 6 (“It is my responsibility to apply feedback to improve my performance”) and 7 (“I hold myself accountable to respond to feedback appropriately”) loaded higher on Utility than on Accountability. However, as this scale originated from an existing and validated questionnaire, and as Cronbach’s alpha of Accountability would not increase when leaving out item 6 and 7, we decided to retain the items in the User Accountability scale. For the Giver Accountability items, in line with the User items, item 21 (“It is my responsibility to give feedback to team members to help them improve their performance”) loaded higher on Giver Utility. Item 23 (“I don’t feel a sense of closure until feedback I have given has been responded to”) loaded higher on User Accountability and, interestingly, was the only Giver item that loaded on a User factor. As removing these items from the Giver Accountability scale would not yield a substantially better reliability and to keep the comparability between the User and Giver scales as good as possible, we decided to continue with the Giver Accountability scale as intended, including the five items (21-25).

**Table 1.** Factor Loadings of DrFOS Items for the 6-factor Solution.

|  | M (SD) | 1 | 2 | 3 | 4 | 5 | 6 |
| --- | --- | --- | --- | --- | --- | --- | --- |
| 1. Feedback contributes to my success in health care practice. | 4.59 (0.57) | .090 | -.007 | **.705** | -.043 | .035 | -.061 |
| 2. To develop my skills in health care practice, I rely on feedback. | 4.67 (0.53) | -.062 | .026 | **.788** | .046 | -.004 | -.017 |
| 3. Feedback is critical for improving performance. | 4.61 (0.58) | .054 | .032 | **.773** | -.040 | -.009 | -.011 |
| 4. Feedback from team members can help me advance in health care practice. | 4.58 (0.57) | .025 | .011 | **.829** | .046 | -.001 | -.034 |
| 5. I find that feedback is critical for reaching my goals. | 4.35 (0.73) | .151 | .004 | **.665** | -.104 | -.003 | .182 |
| 6. It is my responsibility to apply feedback to improve my performance. | 4.32 (0.73) | .064 | -.073 | .251 | .169 | .147 | .180 |
| 7. I hold myself accountable to respond to feedback appropriately. | 4.38 (0.61) | -.047 | -.058 | **.305** | .140 | .195 | .244 |
| 8. I don't feel a sense of closure until I respond to feedback. | 3.16 (0.91) | .027 | .032 | .126 | .021 | -.060 | **.629** |
| 9. If my team member gives me feedback, it is my responsibility to respond to it. | 3.85 (0.84) | -.003 | .038 | .187 | .197 | .030 | **.437** |
| 10. I feel obligated to make changes based on feedback. | 2.89 (0.98) | .094 | -.109 | -.041 | .028 | .009 | **.452** |
| 11. I feel self-assured when dealing with received feedback. | 3.76 (0.75) | -.023 | .041 | .021 | -.052 | **.726** | -.040 |
| 12. Compared to my peers, I am more competent at handling received feedback. | 3.03 (0.71) | .019 | .198 | -.101 | -.194 | **.301** | .228 |
| 13. I believe that I have the ability to deal with received feedback effectively. | 4.06 (0.56) | .034 | .017 | .111 | .150 | **.505** | -.090 |
| 14. I feel confident when responding to both positive and negative feedback. | 3.67 (0.77) | -.014 | .123 | -.028 | -.038 | **.741** | -.034 |
| 15. I know that I can handle the feedback that I receive. | 3.94 (0.66) | .040 | -.053 | .005 | .041 | **.818** | -.011 |
| 16. Me giving feedback to team members contributes to their success in health care practice. | 4.02 (0.67) | **.554** | .052 | .194 | .056 | .020 | -.014 |
| 17. To develop their skills in health care practice, team members rely on my feedback. | 3.70 (0.75) | **.792** | .016 | .043 | -.021 | -.023 | -.027 |
| 18. Me giving feedback is critical for the performance improvement of team members. | 3.65 (0.79) | **.847** | .001 | .041 | -.062 | -.017 | -.019 |
| 19. My feedback can help team members advance in health care practice. | 3.96 (0.67) | **.701** | .043 | .083 | .084 | .064 | -.105 |
| 20. Me giving feedback is critical for team members reaching their goals. | 3.39 (0.86) | **.791** | -.002 | .004 | -.148 | -.038 | .210 |
| 21. It is my responsibility to give feedback to team members to help them improve their performance. | 3.51 (0.84) | **.500** | -.026 | -.162 | **.339** | .107 | .090 |
| 22. I hold myself accountable to give feedback to a team member that can be responded to. | 3.65 (0.83) | .285 | .018 | -.053 | **.488** | .071 | .146 |
| 23. I don't feel a sense of closure until feedback I have given has been responded to. | 2.98 (0.97) | .075 | .125 | -.058 | **.244** | -.160 | **.414** |
| 24. If I give feedback to a team member, it is my responsibility to give feedback that can be responded to. | 3.86 (0.82) | -.013 | .078 | .011 | **.625** | .016 | .163 |
| 25. I feel obligated to give feedback in a way that supports the receiver to make changes based on it. | 4.00 (0.83) | .004 | .031 | .090 | **.604** | .021 | .009 |
| 26. I feel self-assured when giving feedback. | 3.25 (0.83) | .005 | **.729** | .018 | -.002 | .025 | -.057 |
| 27. Compared to peers, I am more competent at giving feedback. | 2.90 (0.77) | .038 | **.654** | -.040 | -.140 | -.048 | .230 |
| 28. I believe that I have the ability to give feedback effectively. | 3.51 (0.74) | .002 | **.714** | .058 | .087 | .093 | -.098 |
| 29. I feel confident that both the positive and negative feedback I give will be responded to. | 3.42 (0.77) | -.032 | **.736** | .021 | .099 | .104 | -.089 |
| 30. I know the feedback that I give can be handled. | 3.76 (0.71) | .210 | **.403** | .004 | .278 | .034 | -.095 |

The estimated reliabilities of six subscale in terms of Cronbach’s alphas ranged from .66 (User Accountability) to .88 (Giver Utility; see Table 2). For none of the items, removing them would lead to a substantial increase in estimated reliability of the scale.

**Table 2.**
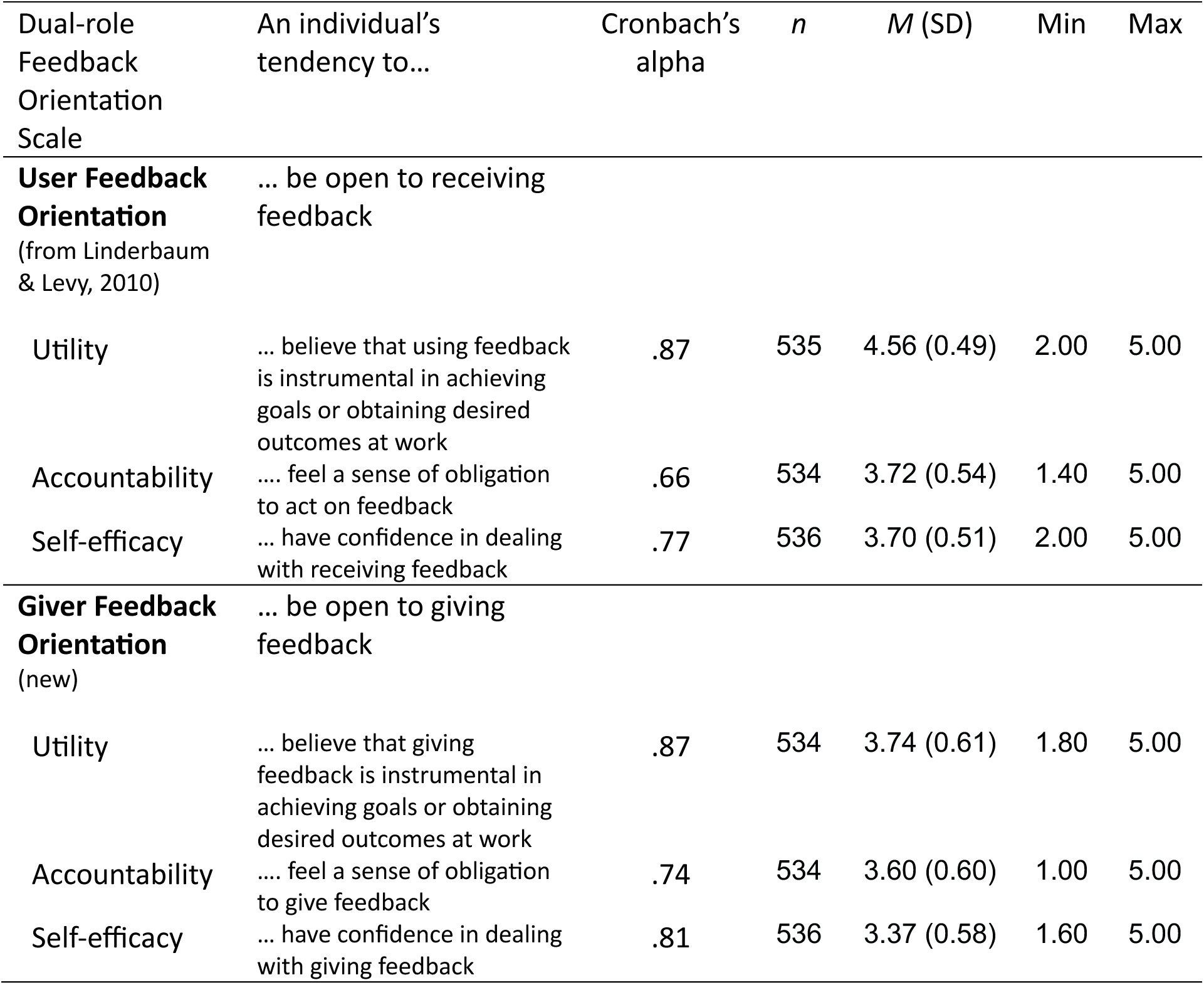
Scale Descriptions, Reliabilities, and Descriptive Statistics.

### Differences and relations between DrFOS subscales

In terms of the scale means, the repeated measures ANOVA showed significant differences between the scales (*F*(5,527.000) = 404.203, *p*<.001, partial η^2^=0.99). Bonferoni posthoc tests indicated that User Utility was significantly higher than all other scales and that Giver Self-efficacy was significantly lower than all other scales (*p*<.001). For User Utility 38% of the participants gave the maximum score of 5 on all five scale items.

### Relations between DrFOS subscales

Table 3 presents the correlations between the six DrFOS subscales. The correlations between the User scales ranged from .15 to .39 and the correlations between Giver scales ranged from .27 to .50. Correlations between the corresponding User and Giver scales were .40, .46, and .44 respectively (bold in Table 3), indicating a range of 16-21% explained variance. This means that even though we see correlations between the corresponding User and Giver scales, the scales do not measure one and the same variable, but more likely represent different constructs.

**Table 3.** Pearson Correlation Coefficients between DrFOS Scales.

|  | **User Feedback Orientation** |  | **Giver Feedback Orientation** |  |  |
| --- | --- | --- | --- | --- | --- |
|  | Accountability | Self-efficacy | Utility | Accountability | Self-efficacy |
| **User Feedback Orientation** |  |  |  |  |  |
| Utility | .39** | .23** | **.40\*\*** | .26** | .07 |
| Accountability | --- | .15** | .39** | **.46\*\*** | .12** |
| Self-efficacy |  | --- | .18** | .16** | **.44\*\*** |
| **Giver Feedback Orientation** |  |  |  |  |  |
| Utility |  |  | --- | .50** | .30** |
| Accountability |  |  |  | --- | .27** |
| Self-efficacy |  |  |  |  | --- |

*Note.* \* $p < .05$ , \*\* $p < .01$ , \*\*\* $P < .001$ .

### Clusters of students

The two-step cluster analysis had a fair cluster quality and yielded three clusters (see Table 4). By far, User Utility was found to have the largest predictor importance. Cluster 1 contained 36% of the participants and can be characterized as relatively high on User Utility and low on all other scales and was therefore labeled “User Utility focused Feedback Orientation”. Cluster 2 contained 35% of the participants and can be characterized by low scores on all scales and was therefore labeled “Low Dual-role Feedback Orientation”. Cluster 3 contained 29% of the participants and can be characterized by relatively high scores on all subscales and was therefore labeled as “High Dual-role Feedback Orientation”. In other words, two clusters have overall low and overall high means on all scales and a third cluster distinguished about a third of the participants having high User Utility, but lower scores on all other scales.

**Table 4.**
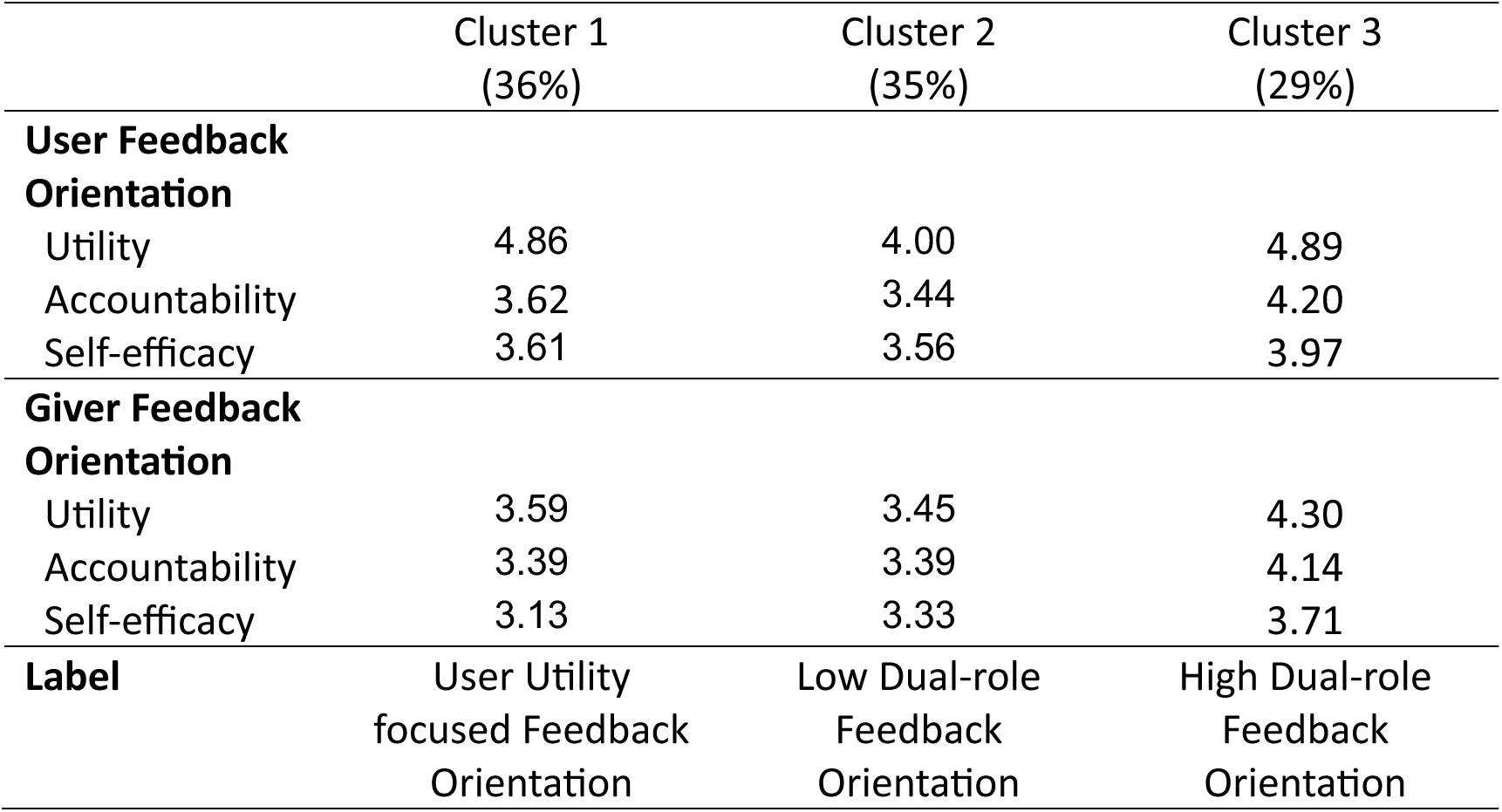
Cluster Means on Subscales from the Two-step Cluster Analysis.

## Discussion

In this study we argued that health professions education should support students’ development as both feedback givers and users. In order to properly address this in research, an instrument is needed to investigate students’ orientations towards receiving and giving feedback. Therefore, we extended the definition of feedback orientation to not only include receptivity to feedback, but also orientation to giving feedback. We mirrored three scales of the Feedback Orientation Scale (Linderbaum & Levy, 2010), and presented and analyzed the Dual-role Feedback Orientation Scale (DrFOS) addressing the research question: to what extent can the DrFOS meaningfully measure and discern giver and user feedback orientations in clinical HPE?

Based on our sample of 537 students, we found that the Giver Feedback Orientation subscales could be meaningfully and reliably discerned from the User subscales. Nearly all subscales were moderately correlated with other scales. Correlations between the mirrored scales were highest: students that valued using feedback, also reported they valued giving feedback; students that felt more accountable for using feedback, also felt more accountable for giving feedback; and students that felt confident they can use feedback, were also more confident that they can give feedback. Overall, the results showed very high scores on *user utility*: students strongly believe their use of the feedback they receive contributes to their professional performance. This might seem obvious and is in line with two studies in medical education (Chen et al., 2022; Mills et al., 2023), but differs from the original findings of Linderbaum and Levy (2010), and two other studies in medical education, who found lower means for students (Rasheed et al., 2015; Thornwal & Ionen, 2024). Though students valued receiving feedback, they did not feel very confident in being able to use it.

Regarding giver feedback orientation, we also found that students rated the value of giving feedback (Giver Utility) higher than their confidence to actually do so. The difference between utility and self-efficacy for both using and giving feedback might indicate a need for additional training in these skills, not only in classroom settings, but also in the complex clinical and interprofessional context. Our cluster analysis showed that one third of the students might not feel the need for such training as they reported high value and high self-efficacy for using and giving feedback. Acknowledging that giving and receiving feedback is not easy or straightforward (Palanganas & Edwards, 2021; Tielemans et al., 2023c), it would be interesting to see whether such training would not only affect students’ self-efficacy but also their utility and accountability.

In a follow-up study, we investigated changes in Dual-role Feedback Orientation, across classroom and workplace phases of training (Tielemans et al., 2025, submitted), and found that students’ lower giver self-efficacy was consistent across training phases. However, their utility as feedback givers significantly dropped in the workplace, suggesting a negative effect of the workplace setting. This research highlighted complexities of the workplace learning environment (e.g., negative modelling behavior, a lack of opportunities to practice giving feedback), identifying specific directions for developing education, and designing educational environments in support of feedback giving. This exemplifies how the Dual-role Feedback Orientation (subdomains) can be used to create specific interventions to bolster proactive feedback processes. In that same follow up study, interprofessional differences are considered, similarly leading to suggestions for the design on more effective learning environments.

### Limitations and Future Research

Future research will need to show whether the instrument yields comparably valid and reliable results. Especially across different context and (health) professions, as our sample was limited to a single institution with only two health professions represented. Second, due to low initial response rates we shortened our questionnaire by excluding the Social Awareness subscale from the original Feedback Orientation Scale. However, as impression management may relate to the complexities students face as they enact feedback skills in the workplace environment, we recommend future studies to include this subscale in the DrFOS. Third, future studies can address the relation between the subscales and other relevant variables such as psychological safety, feedback culture, and educational and patient outcomes. Fourth, longitudinal studies to explore the extent to which user and giver feedback orientations are a rather stable trait or can be affected by experiences and education would be highly relevant. With respect to this, Linderbaum and Levy (2010) themselves indicated that they would expect feedback orientations to be rather stable over time, without targeted interventions. Additionally, though we found that user and giver subscales could be meaningfully and reliably discerned, engaging with questions about using feedback may have affected students’ responses about giving feedback. Future research using Giver Feedback Orientation as a stand-alone concept/instrument, may help further strengthen the DrFOS. Lastly, the relatively low reliability of the user accountability scale is a relevant thing to bear in mind, as well as some items that strictly do not load highest on the intended subscale. Transparently reporting about this in future studies, can aid our understanding of whether specific items can be further improved.

### Conclusion

For (future) health care professionals it is indispensable to recognize the importance of, and practice, both using and giving feedback in the clinical context. The Dual-role Feedback Orientation Scales can be used to reliably and meaningfully distinguish and measure these. Ultimately, this will create opportunities to further sustain and improve students’ giver and user feedback orientations.

## Data Availability

The data that support the findings of this study are not publicly available

## Acknowledgements

The authors would like to thank Zufan Shewangizaw for her help gathering the student data. Furthermore, the authors acknowledge Sjoukje van den Broek, Marieke van der Schaaf, Charlotte Eijkelboom and the LSER PhD students for their contributions to developing the DrFOS instrument. Finally, the authors want to thank Anne Scheel for providing insights and suggestions for the statistical analyses. In loving memory of Tineke Westerveld.

## Tables

DrFOS Items Per Subscale\*
|  | Feedback User | Feedback giver |
| --- | --- | --- |
| Utility | 1. Feedback contributes to my success in health care practice.<br>2. To develop my skills in health care practice, I rely on feedback.<br>3. Feedback is critical for improving performance.<br>4. Feedback from team members can help me advance in health care practice.<br>5. I find that feedback is critical for reaching my goals. | 6. Me giving feedback to team members contributes to their success in health care practice.<br>7. To develop their skills in health care practice, team members rely on my feedback.<br>8. Me giving feedback is critical for the performance improvement of team members.<br>9. My feedback can help team members advance in health care practice.<br>10. Me giving feedback is critical for team members reaching their goals. |
| Accountability | 11. It is my responsibility to apply feedback to improve my performance.<br>12. I hold myself accountable to respond to feedback appropriately.<br>13. I don't feel a sense of closure until I respond to feedback.<br>14. If my team member gives me feedback, it is my responsibility to respond to it.<br>15. I feel obligated to make changes based on feedback. | 16. It is my responsibility to give feedback to team members to help them improve their performance.<br>17. I hold myself accountable to give feedback to a team member that can be responded to.<br>18. I don't feel a sense of closure until feedback I have given has been responded to.<br>19. If I give feedback to a team member, it is my responsibility to give feedback that can be responded to.<br>20. I feel obligated to give feedback in a way that supports the receiver to make changes based on it. |
| Self-efficacy | 21. I feel self-assured when dealing with received feedback.<br>22. Compared to my peers, I am more competent at handling received feedback.<br>23. I believe that I have the ability to deal with received feedback effectively.<br>24. I feel confident when responding to both positive and negative feedback.<br>25. I know that I can handle the feedback that I receive. | 26. I feel self-assured when giving feedback.<br>27. Compared to peers, I am more competent at giving feedback.<br>28. I believe that I have the ability to give feedback effectively.<br>29. I feel confident that both the positive and negative feedback I give will be responded to.<br>30. I know the feedback that I give can be handled. |

\*The excluded Social awareness questions are available upon request, please contact the corresponding author

